## Supplemental for "International travel as a risk factor for carriage of extended-spectrum β-lactamase-producing *Escherichia coli* in a large sample of European individuals - The AWARE Study"

### Supplementary material

Daloha Rodríguez-Molina[1,2,3], Fanny Berglund[4,5], Hetty Blaak[6], Carl-Fredrik Flach[4,5], Merel Kemper[6], Luminita Marutescu[7,8], Gratiela Pircalabioru Gradisteanu[7,8], Marcela Popa[7,8], Beate Spießberger[9,10,11], Laura Wengenroth[1], Mariana Carmen Chifiriuc[7,8], D. G. Joakim Larsson[4,5], Dennis Nowak[1,12], Katja Radon[1], Ana Maria de Roda Husman[6], Andreas Wieser[9,10,11], Heike Schmitt[6].

1: Institute and Clinic for Occupational, Social and Environmental Medicine; University Hospital, LMU Munich, Munich, Germany.

2: Institute for Medical Information Processing, Biometry, and Epidemiology – IBE, LMU Munich, Munich, Germany.

3: Pettenkofer School of Public Health, Munich, Germany.

4: Department of Infectious Diseases, Institute of Biomedicine, The Sahlgrenska Academy, University of Gothenburg, Gothenburg, Sweden.

5: Centre for Antibiotic Resistance Research (CARe), University of Gothenburg, Gothenburg, Sweden.

6: Centre of Infectious Disease Control, National Institute for Public Health and the Environment, Bilthoven, The Netherlands.

7: Department of Microbiology and Immunology, Faculty of Biology, University of Bucharest and the Academy of Romanian Scientists, Bucharest, Romania.

8: Earth, Environmental and Life Sciences Section, Research Institute of the University of Bucharest, University of Bucharest, Bucharest, Romania.

9: German Centre for Infection Research (DZIF) Partner Site Munich.

10: Max von Pettenkofer Institute, Faculty of Medicine, LMU Munich.

11: Department of Infectious Diseases and Tropical Medicine, LMU University Hospital Munich, Munich, Germany.

12: Comprehensive Pneumology Center Munich (CPC-M), German Center for Lung Research (DZL), Munich, Germany.

*Table S1: Multiple imputation diagnostics - Traditional (unweighted) logistic regression models, complete cases vs. imputed for model with personal travel score, AWARE Study, 2022*

| | | **Crude models, OR (95% CI)** | | **Adjusted models, OR (95% CI)** | |
|---|---|---|---|---|---|
| | **Missings** | **Complete cases** | **Imputed, n = 1183** | **Complete cases, n = 991** | **Imputed, n = 1183** |
| Country: Netherlands (ref. = Germany) | 109 | 0.71 (0.41-1.26) | 0.73 (0.42-1.26) | 0.77 (0.43-1.41) | 0.78 (0.44-1.38) |
| Country: Romania (ref. = Germany) | 109 | 1.58 (0.81-3.08) | 1.55 (0.81-2.97) | 1.65 (0.82-3.33) | 1.57 (0.80-3.07) |
| **Educational level: High (ref. = Low)[a]** | 111 | **1.70 (1.02-2.97)** | 1.56 (0.92-2.63) | 1.38 (0.79-2.50) | 1.26 (0.72-2.18) |
| Age (continuous) | 109 | 0.99 (0.97-1.01) | 0.99 (0.98-1.01) | 1.00 (0.98-1.01) | 1.00 (0.98-1.02) |
| Sex: Male (ref. = Female) | 113 | 1.07 (0.67-1.68) | 1.09 (0.70-1.70) | 1.06 (0.65-1.71) | 1.05 (0.66-1.65) |
| **Personal travel score (continuous)[b]** | 112 | **1.32 (1.10-1.65)** | **1.25 (1.09-1.44)** | **1.30 (1.08-1.63)** | **1.24 (1.07-1.43)** |
| Frequency of diarrhea: Often or always (ref. Never, rarely or sometimes) | 109 | 1.61 (0.65-3.44) | 1.53 (0.67-3.52) | 1.66 (0.63-3.76) | 1.55 (0.65-3.71) |
| Antibiotics use: Yes (ref. No) | 121 | 1.12 (0.66-1.86) | 1.10 (0.66-1.82) | 0.96 (0.54-1.64) | 0.95 (0.56-1.61) |

Notes:

[a] Educational level according to the International Standard Classification of Education (ISCED): Low = ISCED 0-2 (Pre-primary education to Lower secondary education), High = ISCED ≥3 (Upper secondary education to Doctoral or equivalent).

[b] Travel score was constructed based on frequency of personal travels to high risk areas for antibiotic resistance in the past year: Includes travels to North Africa, Sub-Saharan Africa, Asia, Central and South America, as well as the European countries Italy, Greece, Bulgaria and Slovenia. The score is the sum of: zero points for not travelling to these areas in the past year, one point for travelling once to these areas in the past year, two points for travelling to these areas two or three times in the past year, and three points for travelling to these areas more than three times in the past year.

Bold highlighting means that the confidence interval does not include the null value.

*Table S2: Multiple imputation diagnostics - Traditional (unweighted) logistic regression models, complete cases vs. imputed for model with individual travel areas, AWARE Study, 2022*

| | | **Crude models, OR (95% CI)** | | **Adjusted models, OR (95% CI)** | |
|---|---|---|---|---|---|
| | **Missings** | **Complete cases** | **Imputed, n = 1183** | **Complete cases, n = 992** | **Imputed, n = 1183** |
| Country: Netherlands (ref. = Germany) | 109 | 0.71 (0.41-1.26) | 0.73 (0.42-1.26) | 0.70 (0.39-1.31) | 0.71 (0.39-1.29) |
| Country: Romania (ref. = Germany) | 109 | 1.58 (0.81-3.08) | 1.55 (0.81-2.97) | 1.70 (0.78-3.67) | 1.71 (0.82-3.55) |
| **Educational level: High (ref. = Low)[a]** | 111 | **1.70 (1.02-2.97)** | 1.56 (0.92-2.63) | 1.27 (0.71-2.35) | 1.19 (0.67-2.13) |
| Age (continuous) | 109 | 0.99 (0.97-1.01) | 0.99 (0.98-1.01) | 1.00 (0.98-1.01) | 1.00 (0.98-1.02) |
| Sex: Male (ref. = Female) | 113 | 1.07 (0.67-1.68) | 1.09 (0.70-1.70) | 0.92 (0.56-1.51) | 0.99 (0.62-1.58) |
| Travels to Europe in the past year: At least once (ref. = Never) | 112 | 0.74 (0.46-1.22) | 0.76 (0.47-1.23) | 0.73 (0.42-1.27) | 0.78 (0.46-1.32) |
| **Travels to Sub-Saharan Africa in the past year: At least once (ref. = Never)** | 109 | **3.99 (1.42-9.69)** | **4.02 (1.64-9.89)** | 2.74 (0.71-8.29) | **4.14 (1.50-11.44)** |
| **Travels to Northern Africa in the past year: At least once (ref. = Never)** | 114 | **3.95 (1.79-8.04)** | **3.72 (1.78-7.78)** | **4.31 (1.84-9.41)** | **3.80 (1.66-8.68)** |
| **Travels to Asia in the past year: At least once (ref. = Never)** | 114 | **3.87 (2.06-6.97)** | **3.73 (2.03-6.86)** | **3.56 (1.77-6.86)** | **3.46 (1.74-6.87)** |
| **Travels to North America in the past year: At least once (ref. = Never)** | 115 | **2.87 (1.13-6.37)** | **2.61 (1.10-6.23)** | 2.77 (0.91-7.24) | 2.40 (0.89-6.45) |
| Travels to Central or South America in the past year: At least once (ref. = Never) | 113 | 0.69 (0.11-2.31) | 0.93 (0.26-3.28) | 0.19 (0.01-1.24) | 0.40 (0.07-2.45) |
| Travels to Australia and Oceania in the past year: At least once (ref. = Never) | 113 | 1.26 (0.07-6.68) | 1.82 (0.24-13.88) | 0.33 (0.01-2.60) | 0.20 (0.01-4.22) |
| Frequency of diarrhea: Often or always (ref. Never, rarely or sometimes) | 115 | 1.61 (0.65-3.44) | 1.53 (0.67-3.52) | 1.36 (0.51-3.15) | 1.25 (0.50-3.12) |
| Antibiotics use: Yes (ref. No) | 115 | 1.12 (0.66-1.86) | 1.10 (0.66-1.82) | 0.89 (0.49-1.55) | 0.91 (0.53-1.55) |

Notes:

[a] Educational level according to the International Standard Classification of Education (ISCED): Low = ISCED 0-2 (Pre-primary education to Lower secondary education), High = ISCED ≥3 (Upper secondary education to Doctoral or equivalent).

Bold highlighting means that the confidence interval does not include the null value.

*Table S3: Models comparing risk factors for ESBL-producing E. coli in stool samples, with personal travel score, AWARE Study, 2022*

| | **Unweighted cOR (95% CI)[a]** | **Unweighted aOR (95% CI)[b]** | **IPW cOR (95% CI)[a,c]** | **IPW aOR (95% CI)[b,c]** |
|---|---|---|---|---|
| Country: Netherlands (ref. = Germany) | 0.73 (0.42-1.26) | 0.78 (0.44-1.38) | 0.73 (0.42-1.26) | 0.75 (0.42-1.33) |
| Country: Romania (ref. = Germany) | 1.55 (0.81-2.97) | 1.57 (0.80-3.07) | 1.55 (0.81-2.97) | 1.61 (0.81-3.21) |
| Educational level: High (ref. = Low)[d] | 1.56 (0.92-2.63) | 1.26 (0.72-2.18) | 1.43 (0.83-2.47) | 1.24 (0.70-2.18) |
| Age (continuous) | 0.99 (0.98-1.01) | 1.00 (0.98-1.02) | 1.00 (0.98-1.01) | 1.00 (0.98-1.02) |
| Sex: Male (ref. = Female)[e] | 1.09 (0.70-1.70) | 1.05 (0.66-1.65) | 1.08 (0.65-1.77) | 1.04 (0.62-1.74) |
| Frequency of diarrhea: Often or always (ref. Never, rarely or sometimes) | 1.53 (0.67-3.52) | 1.55 (0.65-3.71) | 1.73 (0.75-4.01) | 1.63 (0.62-4.26) |
| Antibiotics use: Yes (ref. No) | 1.10 (0.66-1.82) | 0.95 (0.56-1.61) | 0.89 (0.49-1.61) | 0.80 (0.42-1.51) |
| **Personal travel score (continuous)** | **1.25 (1.09-1.44)** | **1.24 (1.07-1.43)** | **1.31 (1.03-1.66)** | **1.28 (1.01-1.64)** |

Notes:
[a]cOR: crude odds ratio.
[b]aOR: adjusted odds ratio.
[,c]IPW: Inverse Probability Weighted model.
[d]Educational level according to the International Standard Classification of Education (ISCED): Low = ISCED 0-2 (Pre-primary education to Lower secondary education), High = ISCED ≥3 (Upper secondary education to Doctoral or equivalent).
[e] Travel score was constructed based on frequency of personal travels to high risk areas for antibiotic resistance in the past year: Includes travels to North Africa, Sub-Saharan Africa, Asia, Central and South America, as well as the European countries Italy, Greece, Bulgaria and Slovenia. The score is the sum of: zero points for not travelling to these areas in the past year, one point for travelling once to these areas in the past year, two points for travelling to these areas two or three times in the past year, and three points for travelling to these areas more than three times in the past year.
ESBL: Extended-Spectrum Beta-Lactamases.
AR: Antibiotic Resistance.
Bold highlighting means that the confidence interval does not include the null value.

*Table S4: Models comparing risk factors for ESBL-producing E. coli in stool samples, with individual travel areas, AWARE Study, 2022*

| | **Unweighted cOR (95% CI)[a]** | **Unweighted aOR (95% CI)[b]** | **IPW cOR (95% CI)[a,c]** | **IPW aOR (95% CI)[b,c]** |
|---|---|---|---|---|
| Country: Netherlands (ref. = Germany) | 0.73 (0.42-1.26) | 0.71 (0.39-1.29) | 0.73 (0.42-1.26) | 0.68 (0.37-1.24) |
| Country: Romania (ref. = Germany) | 1.55 (0.81-2.97) | 1.71 (0.82-3.55) | 1.55 (0.81-2.97) | 1.85 (0.87-3.94) |
| Educational level: High (ref. = Low)[d] | 1.56 (0.92-2.63) | 1.19 (0.67-2.13) | 1.43 (0.83-2.47) | 1.16 (0.64-2.08) |
| Age (continuous) | 0.99 (0.98-1.01) | 1.00 (0.98-1.02) | 1.00 (0.98-1.01) | 1.00 (0.98-1.02) |
| Sex: Male (ref. = Female) | 1.09 (0.70-1.70) | 0.99 (0.62-1.58) | 1.08 (0.65-1.77) | 0.96 (0.58-1.61) |
| Frequency of diarrhea: Often or always (ref. Never, rarely or sometimes) | 1.53 (0.67-3.52) | 1.25 (0.50-3.12) | 1.73 (0.75-4.01) | 1.29 (0.46-3.58) |
| Antibiotics use: Yes (ref. No) | 1.10 (0.66-1.82) | 0.91 (0.53-1.55) | 0.89 (0.49-1.61) | 0.74 (0.39-1.41) |
| Travels to Europe in the past year: At least once (ref. = Never) | 0.76 (0.47-1.23) | 0.78 (0.46-1.32) | 0.90 (0.52-1.56) | 0.83 (0.46-1.48) |
| **Travels to Sub-Saharan Africa in the past year: At least once (ref. = Never)** | **4.02 (1.64-9.89)** | **4.14 (1.50-11.44)** | **4.83 (1.96-11.91)** | **4.60 (1.60-13.26)** |
| **Travels to Northern Africa in the past year: At least once (ref. = Never)** | **3.72 (1.78-7.78)** | **3.80 (1.66-8.68)** | **3.79 (1.74-8.23)** | **4.03 (1.67-9.68)** |
| **Travels to Asia in the past year: At least once (ref. = Never)** | **3.73 (2.03-6.86)** | **3.46 (1.74-6.87)** | **4.44 (2.36-8.35)** | **4.08 (1.97-8.43)** |
| **Travels to North America in the past year: At least once (ref. = Never)** | **2.61 (1.10-6.23)** | 2.40 (0.89-6.45) | **2.79 (1.17-6.67)** | 2.40 (0.94-6.09) |
| Travels to Central or South America in the past year: At least once (ref. = Never) | 0.93 (0.26-3.28) | 0.40 (0.07-2.45) | 0.78 (0.21-2.95) | 0.33 (0.05-2.15) |
| Travels to Australia and Oceania in the past year: At least once (ref. = Never) | 1.82 (0.24-13.88) | 0.20 (0.01-4.22) | 1.70 (0.19-14.83) | 0.19 (0.01-7.09) |

Notes:
[a]cOR: crude odds ratio.
[b]aOR: adjusted odds ratio.
[,c]IPW: Inverse Probability Weighted model.
[d]Educational level according to the International Standard Classification of Education (ISCED): Low = ISCED 0-2 (Pre-primary education to Lower secondary education), High = ISCED ≥3 (Upper secondary education to Doctoral or equivalent).
ESBL: Extended-Spectrum Beta-Lactamases.
AR: Antibiotic Resistance.
Bold highlighting means that the confidence interval does not include the null value.
